# Evaluating the Interplay and Impact of Quality and Technical Variations on Performance of AI Models in Multi-Institutional Prostate MRI Datasets

**DOI:** 10.64898/2026.01.19.26344376

**Authors:** Sena Azamat, Amritpal Singh, Abhishek Midya, Zelin Zhang, Michael Gilkey, Farid Hajibonabi, Pranav Sompalle, Ardeshir R. Rastinehad, Sadhna Verma, Elias George Kikano, Martin Sanda, Mohammad Hajiha, Rakesh Shiradkar, Satish E. Viswanath, Anant Madabhushi

## Abstract

**Purpose:** To evaluate whether visually assessed high-quality prostate MRI, as defined by the Prostate Imaging Quality (PI-QUAL) scoring system, is sufficient to ensure consistent AI model performance, or whether hidden acquisition-related technical effects introduce variability that cannot be detected through visual inspection.

**Methods:** This retrospective study included 568 bpMRI scans from seven institutions, classified using PI-QUAL v2 into high-quality (scores 2-3) and low-quality (score 1) groups. Quantitative image quality features were extracted using MRQy from T2w and ADC sequences. Three experiments were conducted on high-quality scans: (1) unsupervised clustering to identify hidden technical batch effects; (2) V-Net models (M₁) for prostate capsule segmentation on T2w using single-cluster and balanced multi-cluster training strategies; and (3) VoxelMorph-based deformable registration (M₂) to align ADC maps to T2w images. Performance metrics included DSC, precision, recall, volume ratio, HD95, ASSD, and TRE. Kruskal-Wallis tests assessed cross-cluster variability, Two One-Sided Tests (TOST) evaluated performance equivalence, and bootstrap resampling quantified training efficiency.

**Results:** Clustering identified four clusters among 277 high-quality bpMRI scans. Single-cluster models showed variable performance (DSC=0.41-0.85) with significant batch effects (p<0.001), even with 140 training samples. The balanced multi-cluster model (n=60) eliminated batch effects (p=0.075), achieved equivalent performance (p<0.001), and demonstrated 2.23-fold greater training efficiency despite using 2.3-fold fewer samples. Registration performance also varied significantly across clusters (TRE=0.86-5.47 mm, p<0.001).

**Conclusion:** Acquisition-related batch effects significantly impact AI generalizability in prostate MRI, even among high-quality scans. Quantitative clustering enables detection of technical variations and supports diversity-aware training strategies that achieve equivalent performance with 2.3-fold fewer samples while eliminating batch effects.

**Summary:** Acquisition-related batch effects significantly impair AI generalizability in prostate MRI, even among scans meeting PI-QUAL quality standards. Clustering-based diversity-aware training strategies successfully eliminated these batch effects and achieved 2.23-fold greater training efficiency compared to single-site approaches, demonstrating that strategic technical diversity outweighs dataset size for developing robust, clinically deployable AI models.

**Key Points:**

- Acquisition-related batch effects significantly impair AI performance on prostate MRI, exceeding the impact of visual image quality as assessed by PI-QUAL scoring.
- Even the model trained on the largest single cluster exhibited significant batch effects (p<0.001). In contrast, a balanced multi-cluster model eliminated cross-cluster variability (p=0.075) while maintaining equivalent performance and requiring a smaller sample size.
- Diversity-aware training achieved 2.23-fold greater training efficiency (95% CI: 2.18-2.29) with 2.3-fold fewer samples, demonstrating that technical diversity outweighs data quantity for robust AI generalization.

**Clinical relevance:** Addressing batch effects is essential for developing reliable, generalizable AI tools for prostate MRI interpretation.

## Introduction

Prostate cancer (PCa) is the second leading cause of cancer-related death among men worldwide (1, 2). Multiparametric prostate MRI (mpMRI) is widely utilized for early PCa detection and is endorsed by international guidelines (3). However, its widespread adoption has also introduced challenges in achieving standardized interpretation. To address this, the Prostate Imaging Reporting and Data System (PIRADS) was developed with support from European and American urology associations (4). PIRADS standardizes MRI acquisition and interpretation (5), with detection rates for clinically significant PCa rising from <10% in PIRADS 1 –2 to >80% in PIRADS 5 (6); however, studies have shown that image quality remains a critical determinant of accuracy, as suboptimal acquisition parameters can obscure subtle lesion characteristics and heighten inter-reader variability among radiologists (7). To address these shortcomings, the Prostate Imaging Quality (PI-QUAL) system was introduced to assess the technical adequacy of scans and guide decisions on repeat imaging and interpretation confidence, particularly for intermediate-risk category (8). While PIRADS and PI-QUAL primarily focus on radiologic workflows, recent advances in artificial intelligence (AI) are transforming prostate MRI interpretation (9). AI models have achieved expert-level performance in controlled settings, but their generalizability in real-world practice remains limited. A key barrier is the issue of batch effects, a term used to refer to non-biological technical variations introduced by differences in scanners, acquisition protocols, institutions, or time points (10, 11). Unlike radiologists, who rely on anatomical and biological context such as comparing lesion signal intensity to reference tissues (12), AI models typically tend to process images as arrays of numerical pixel values, learning statistical patterns that are typically devoid of a deep understanding of underlying anatomy or biology (13). As a result, AI systems are especially vulnerable to batch effects, even when scans appear visually similar (14, 15). This vulnerability often leads to substantial performance drops on external datasets compared with training data (15, 16), underscoring the need for robust strategies to identify and mitigate acquisition-related variability.

Several tools have been developed over the years to assess batch effects in the medical imaging domain, including Batch Effect Explorer (BEEx) and MRQy (11, 17). While BEEx supports both radiology and pathology images, MRQy is a previously developed image quality tool optimized for MRI and offers a more extensive set of modality-specific quality metrics (17). These include intensity-based features (e.g., mean and standard deviation of voxel intensities), entropy (image complexity), signal-to-noise ratio (SNR), and coefficient of joint variation (uniformity). MRQy also extracts metadata such as voxel resolution and matrix size, critical indicators of acquisition variability. By quantifying these metrics, MRQy enables unsupervised clustering to identify site or scanner specific patterns not discernible through visual inspection. This facilitates early detection and mitigation of batch effects prior to downstream model development. For MRI datasets, MRQy offers advantages of specificity, comprehensive set of metrics, and ability to generalize across anatomical sites.

In this study, we asked whether visually assessed high-quality MR imaging, as curated by the PI-QUAL scoring system, is sufficient to ensure consistent AI model performance across different institutions. A corollary to this question was whether hidden acquisition-related technical batch effects introduce variability in AI model performance that cannot be detected through visual inspection alone. Although PI-QUAL was initially developed for multiparametric MRI, it has also been applied to biparametric protocols, which exclude dynamic contrast-enhanced sequences. We selected biparametric prostate MRI (bpMRI) for this study due to its growing clinical adoption and relevance to AI applications (18). To investigate this question, we focused on two key AI tasks: prostate gland segmentation and deformable multi-sequence image registration. Our hypothesis for this study was that if visual quality were truly sufficient for developing robust AI models, the subsequent AI models would achieve stable and consistent performance across sites. Conversely, if subtle technical variations exert a stronger influence, performance degradation on external datasets would reflect the fact that visual assessment alone does not capture the full extent of acquisition variability.

## Materials and Methods

### Dataset

A total of 568 biparametric MRI scans from seven centers were retrospectively analyzed, including 409 in-house cases and 159 from a public database (Table 1). Eligible scans were required to contain tri-planar T2-weighted (T2w) images (acquired in axial, sagittal, and coronal planes), high b-value diffusion-weighted imaging (DWI) with maximum b-value ≥1400 s/mm², and apparent diffusion coefficient (ADC) maps calculated from at least two b-values (b=0 and b=800-1000 s/mm²). Cases with incomplete sequence coverage, corrupted DICOM files, or longitudinal follow-up imaging were excluded (Figure 1). The workflow (Figure 2) included three stages: (1) visual quality assessment using PI-QUAL v2 and extraction of MRQy-derived image quality metrics from T2w and ADC sequences, (2) unsupervised clustering of MRQy features using Uniform Manifold Approximation and Projection (UMAP) to identify site-specific patterns, and (3) cross-cluster evaluation of AI models for prostate capsule segmentation on T2w images and deformable multi-sequence registration of ADC maps onto T2w images. These tasks were selected as foundational components of prostate MRI analysis, as segmentation enables prostate volume estimation and treatment planning, while registration is essential for building computer-aided detection algorithms (19–21).

**Figure 1.**
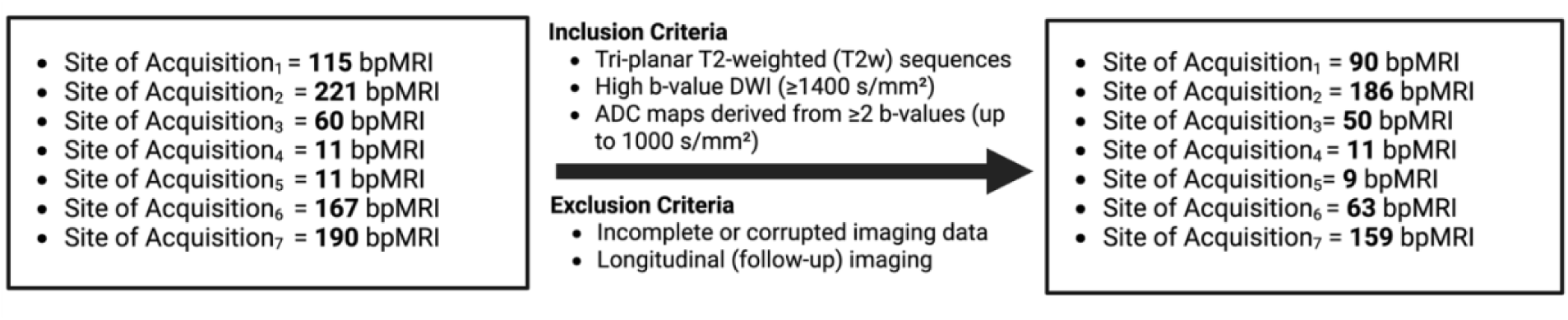
Inclusion and Exclusion Criteria for Multi-center Biparametric MRI Analysis.

**Figure 2.**
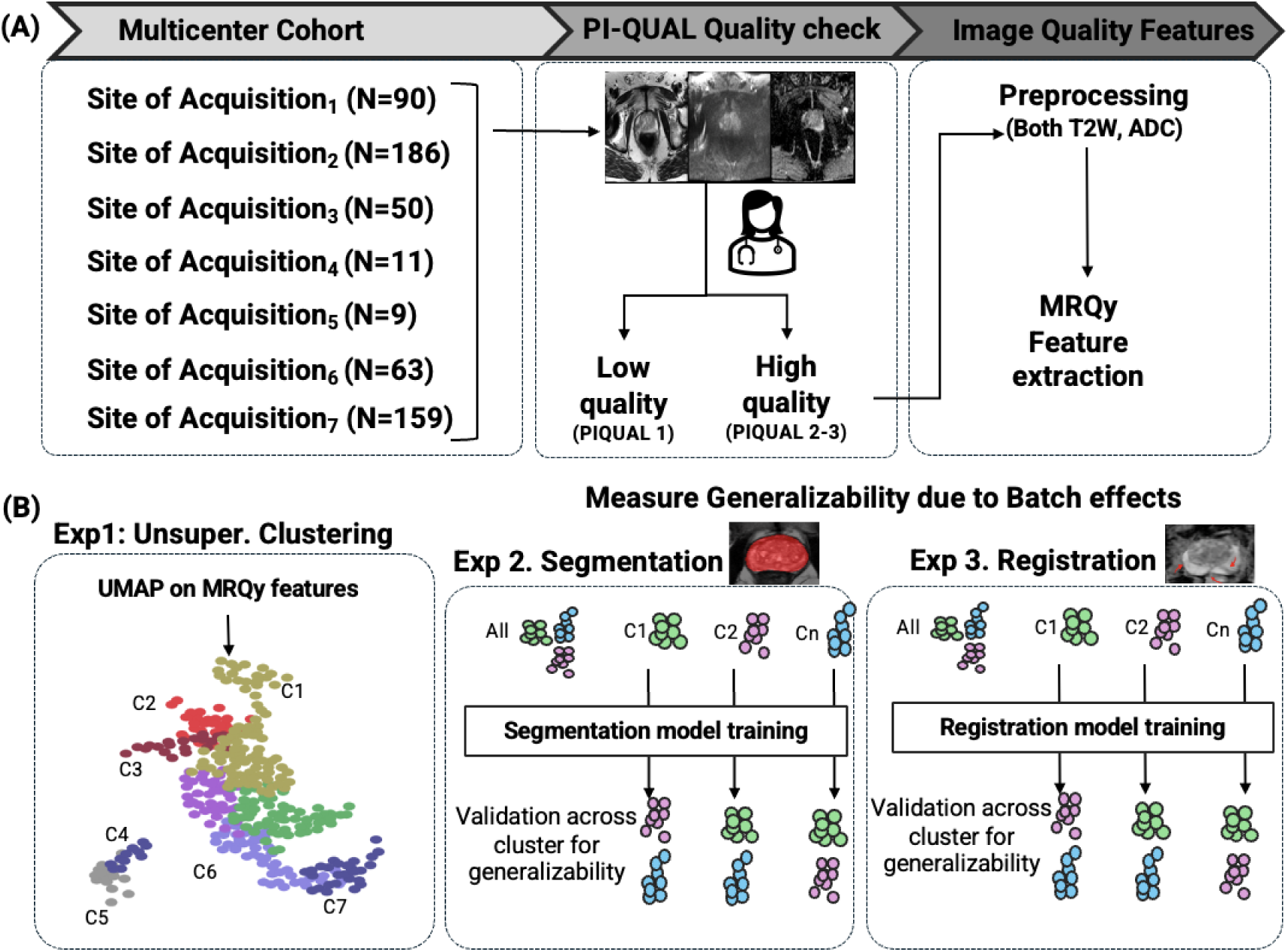
Overview of the study workflow for analyzing multi-site batch effects in prostate MRI. (A) Multi-center cohort data collection from seven institutions (N=568 total), followed by PIQUAL quality assessment and MRQy feature extraction from both T2W and ADC sequences. (B) Three-part generalizability analysis was conducted: Experiment 1 shows unsupervised clustering of MRQy features using UMAP to identify site-specific clusters; Experiments 2 and 3 were focused on evaluating cross-site model performance for prostate gland segmentation and deformable multi-sequence image registration tasks, respectively, with validation across different institutional clusters to assess the impact of protocol heterogeneity on AI model generalizability.

**Table 1.** Distribution of High-Quality (PI-QUAL 2-3) bpMRI Scans (n=277) across Seven Institutions and Four Technical Clusters.

| Acquisition Site (Total Number) | Cluster 1 | Cluster 2 | Cluster 3 | Cluster 4 |
| --- | --- | --- | --- | --- |
| Site of Acquisition<sub>1</sub> (N=90) | 50 | 0 | 0 | 2 |
| Site of Acquisition<sub>2</sub> (N=186) | 10 | 49 | 0 | 1 |
| Site of Acquisition<sub>3</sub> (N=50) | 0 | 0 | 42 | 0 |
| Site of Acquisition<sub>4</sub> (N=11) | 0 | 0 | 0 | 11 |
| Site of Acquisition<sub>5</sub> (N=9) | 9 | 0 | 0 | 0 |
| Site of Acquisition<sub>6</sub> (N=63) | 0 | 0 | 2 | 30 |
| Site of Acquisition<sub>7</sub> (N=159) | 71 | 0 | 0 | 0 |
| Total per Cluster | 140 | 49 | 44 | 44 |

### Evaluation of Image Quality According to PI-QUAL v2

Image quality assessment was performed according to PI-QUAL v2 criteria (8), where the detailed definitions are provided. Tri-planar T2w, high b-value DWI, and ADC maps were independently evaluated for signal-to-noise ratio, anatomical delineation, contrast, and absence of artifacts, with scores assigned per sequence. Each bpMRI was classified as inadequate (score 1), acceptable (score 2), or optimal (score 3), and subsequently grouped into low quality (scores 1) or high quality (score 2-3). Low-quality scans were excluded from subsequent analyses to isolate technical batch effects from overt quality deficiencies. Including these inadequate scans would confound performance variations with obvious quality problems, obscuring the hidden acquisition-related factors that persist even within quality-compliant data.

### Experiment 1. Unsupervised Clustering of Image Quality Metrics

Image quality metrics were extracted from axial T2w images and ADC maps using MRQy, which quantifies acquisition-related characteristics directly from raw image volumes without predefined regions of interest. For each scan, T2w and ADC metrics were merged and standardized using z-score normalization. Unsupervised clustering was then performed exclusively on the PI-QUAL-defined high-quality group to investigate whether hidden technical batch effects persist even within visually homogeneous, high-quality scans. Dimensionality reduction was carried out with Uniform Manifold Approximation and Projection (UMAP), followed by k-means clustering to reveal site-specific acquisition patterns (17, 22). Cluster overlap accuracy was calculated by labeling each cluster with its dominant institution and computing the proportion of correctly grouped scans. We hypothesized that site-specific acquisition patterns would be reflected as distinct clusters even within the PI-QUAL-defined high-quality subset, indicating that visual quality assessment alone is insufficient to capture all acquisition-related technical variations.

### Experiment 2. Impact on generalizability of prostate gland segmentation due to batch effects

To evaluate the impact of batch effects on prostate gland segmentation, we trained and tested a V-Net deep learning model (M_1_) on T2w MRI scans, using the MRQy-defined clusters within the high PI-QUAL group from Experiment 1 (23). Ground truth prostate gland segmentations were manually annotated by a board-certified radiologist (xx) with 9 years of experience in genitourinary imaging, interpreting over 1,500 prostate MRI examinations annually. Two complementary experiments were designed. In the first, the M_1_ model was trained individually on each cluster and then evaluated on the remaining clusters. This setup allowed for quantifying the generalization gap introduced by training on site-specific data. In the second experiment, a balanced training set was created by randomly selecting 15 cases from each cluster. The model trained on this multi-cluster subset was then evaluated on the remaining, non-overlapping data from all clusters (Figure 3). This design allowed for control of class imbalance to isolate the effect of acquisition variability on model performance. The same preprocessing pipeline and model architecture were applied in both experiments. Preprocessing included normalization, binarization of segmentation masks, and data augmentation; full details are provided in the Supplementary Material. Each model was trained for up to 100 epochs, and the best-performing version based on validation Dice similarity coefficient (DSC) was retained. Performance was measured using DSC, precision, recall, Intersection over Union (IoU), volume ratio, Hausdorff Distance at the 95th percentile (HD95), and Average Symmetric Surface Distance (ASSD). Statistical comparisons across clusters were performed using the Kruskal-Wallis test, and p<0.05 was accepted as statistically significant.

**Figure 3.**
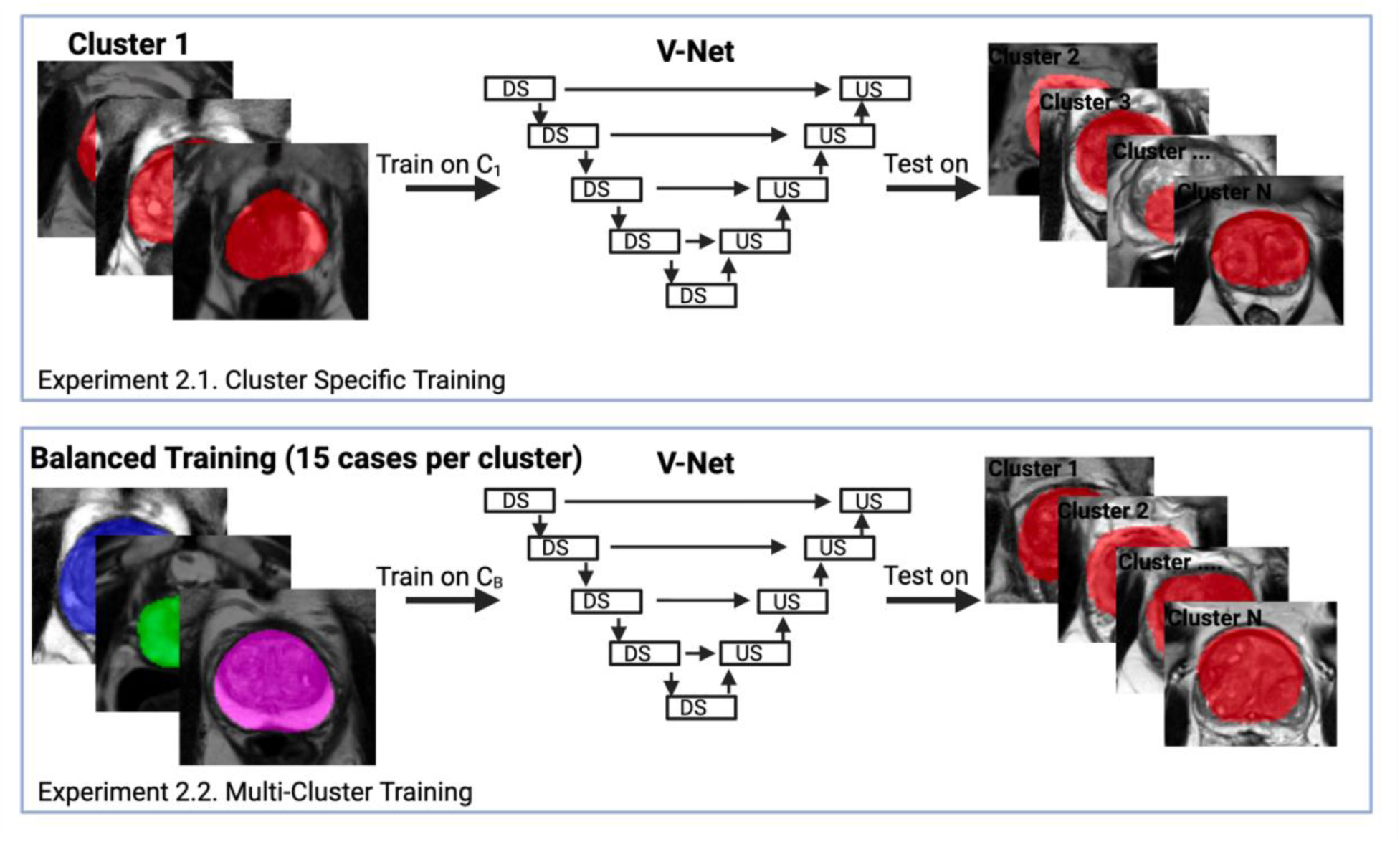
Experimental design for assessing the impact of batch effects on prostate gland segmentation. Two complementary experiments were performed using a V-Net model (M_1_). Top (Cluster-Specific Training) The model was trained on individual MRQy-defined clusters and evaluated on the remaining clusters to quantify the generalization gap introduced by site-specific training. Bottom (Balanced Multi-Cluster Training) 15 cases were randomly selected from each cluster to create a balanced training set. The model was then evaluated on the remaining, non-overlapping data to isolate the effect of acquisition variability while controlling for class imbalance. Both experiments used the same model architecture and evaluation metrics, including Dice Similarity Coefficient (DSC), precision, recall, Intersection over Union (IoU), volume ratio, Hausdorff Distance at the 95th percentile (HD95), and Average Symmetric Surface Distance (ASSD). Statistical comparisons were conducted using the Kruskal–Wallis test. DS = downsampling; US = upsampling.

In addition to cross-cluster performance evaluation, we conducted two analyses to compare training approaches. First, equivalence testing using the Two One-Sided Tests (TOST) procedure assessed whether the multi-cluster model achieved comparable segmentation performance to the best single-cluster model. An equivalence margin of ±0.10 DSC was pre-specified based on typical inter-rater variability in prostate gland segmentation (24). Statistical equivalence was concluded when the 90% confidence interval for the mean difference fell entirely within equivalence bounds (p<0.05). Second, we quantified training efficiency as the mean DSC divided by the number of training samples. Bootstrap resampling with replacement was used to construct 95% confidence intervals for the efficiency ratio between approaches. A confidence interval excluding 1.0 indicated statistically significant differences in training efficiency.

### Experiment 3. Evaluating generalizability of registration algorithm due to batch effects

To evaluate the impact of batch effects on deformable image registration, we trained and tested a VoxelMorph-based registration (25) model (M_2_) using the MRQy-defined clusters among high PI-QUAL group from Experiment 1. In each case, the T2w image served as the fixed reference due to its superior spatial resolution, while the ADC map was treated as the moving image. Preprocessing steps, hierarchical rigid and affine alignment, and training details are provided in the Supplementary Material. Model development employed a five-fold cross-validation, followed by retraining of the best configuration on the full dataset of high-quality scans (Figure 4). Performance was assessed using DSC for prostate capsule overlap and target registration error (TRE) for center-based spatial alignment. Statistical comparisons across clusters were performed using the Kruskal–Wallis test.

**Figure 4.**
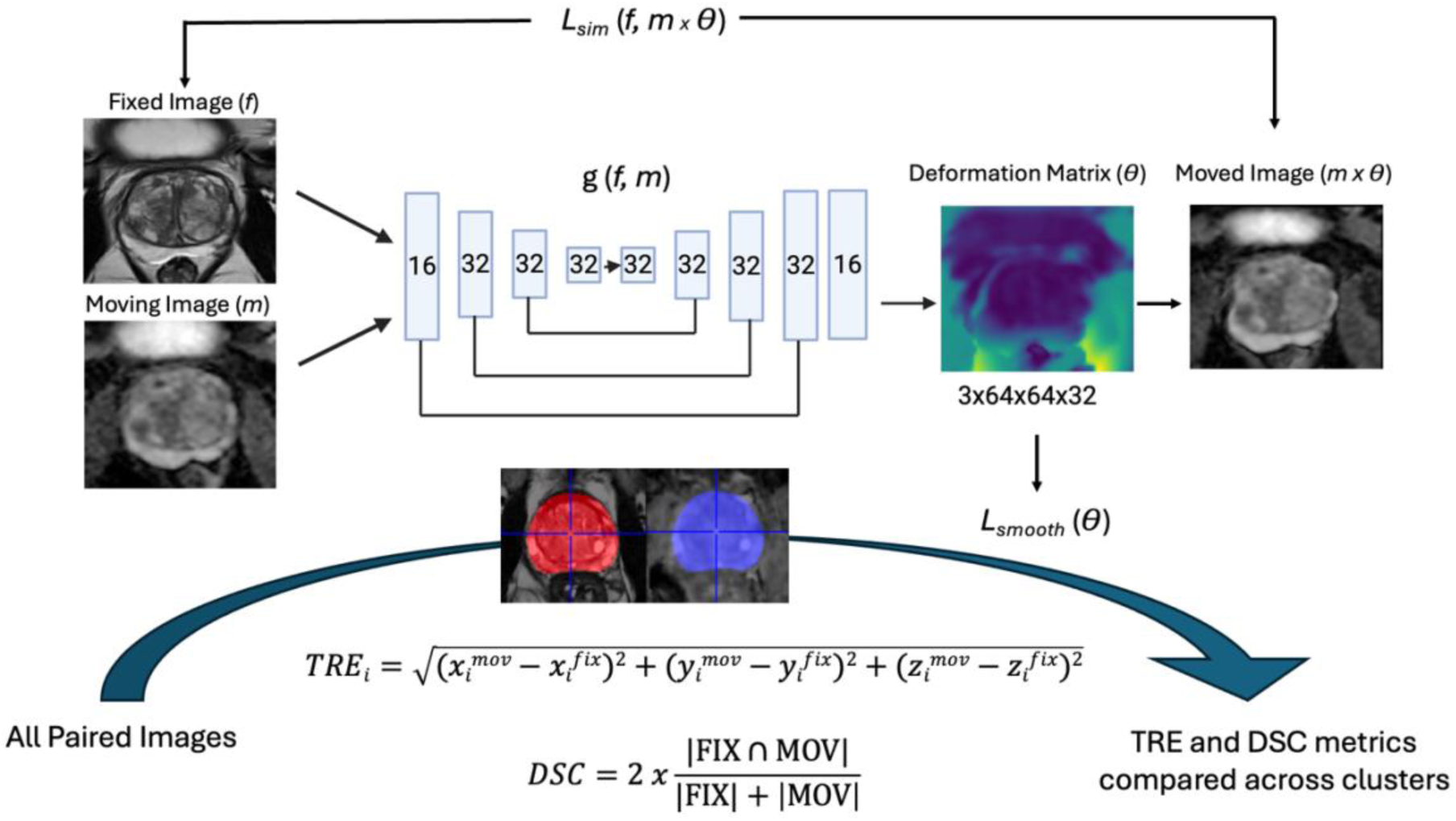
The architecture takes a fixed T2w MRI (f) and a moving ADC map (m) as inputs. These images are processed through the function g(f, m), which represents a U-Net backbone featuring encoder-decoder paths. The network outputs a deformation matrix (θ) of size 3×64×64×32, which is applied to the moving image through a spatial transformation to produce the moved image (m×θ). The network is trained using two loss functions: a similarity loss *Lsim* (f, m×θ) that measures alignment between the fixed and moved images, and a smoothness loss *Lsmooth* (θ) that ensures deformation field regularity. The registration quality is evaluated using Target Registration Error (TRE) calculated as the Euclidean distance between corresponding landmarks in registered and ground truth images, and Dice Similarity Coefficient (DSC) measuring overlap between prostatic gland segmentation on moved and fixed images. These metrics are compared across different image clusters to assess registration performance.

**Figure 5.**
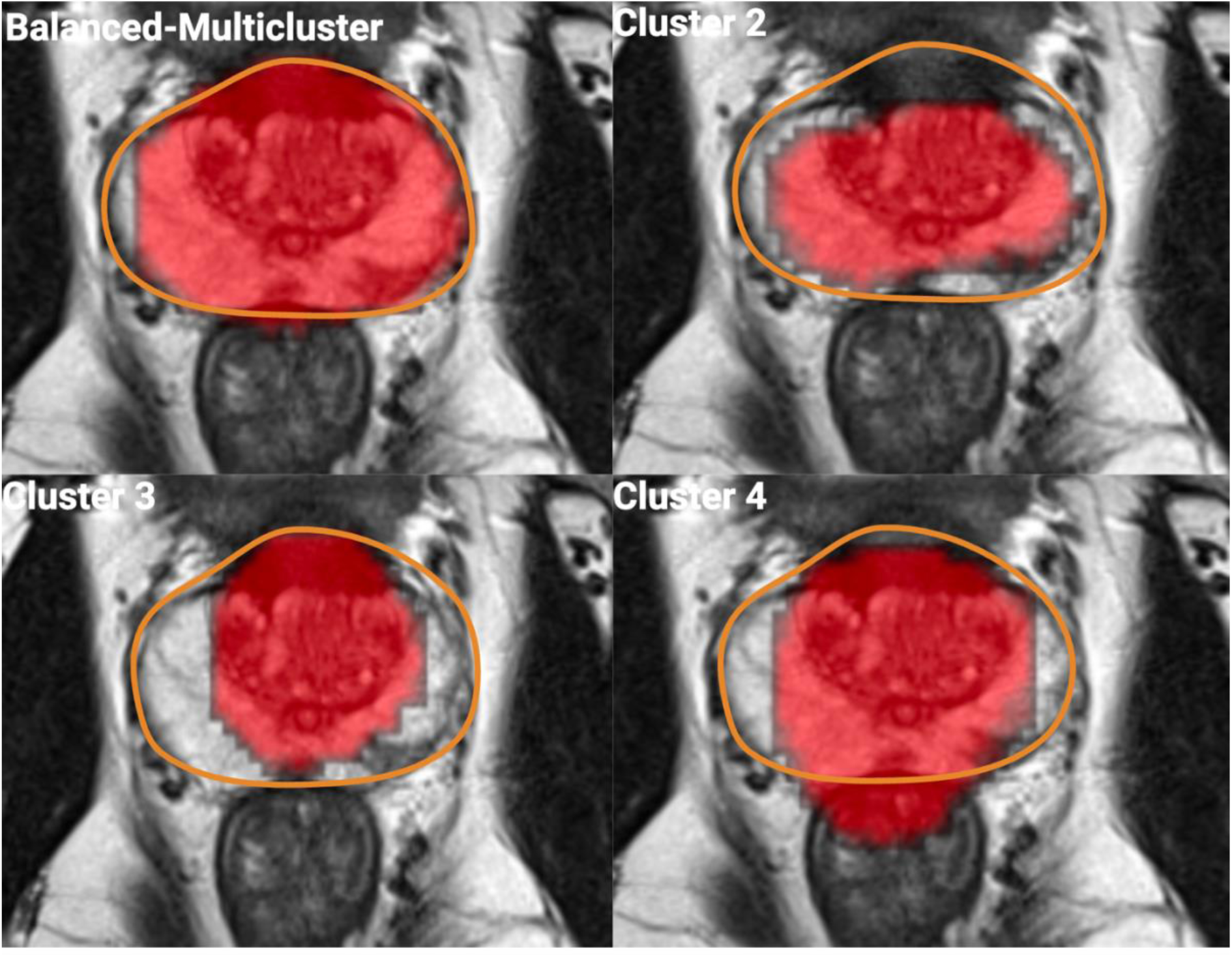
Qualitative comparison of prostate gland segmentation results across different training approaches. Representative axial T2-weighted MRI slices showing predicted segmentation masks (red overlay) generated by models trained on individual clusters (C_2-4_; the test case was selected from C_1_) versus the balanced multi-cluster approach. The balanced multi-cluster model demonstrates superior segmentation quality with well-defined boundaries that closely match the ground truth, while single-cluster models show varying degrees of over and under segmentations (C_2_ to C_4_) or boundary irregularitie.

**Figure 6.**
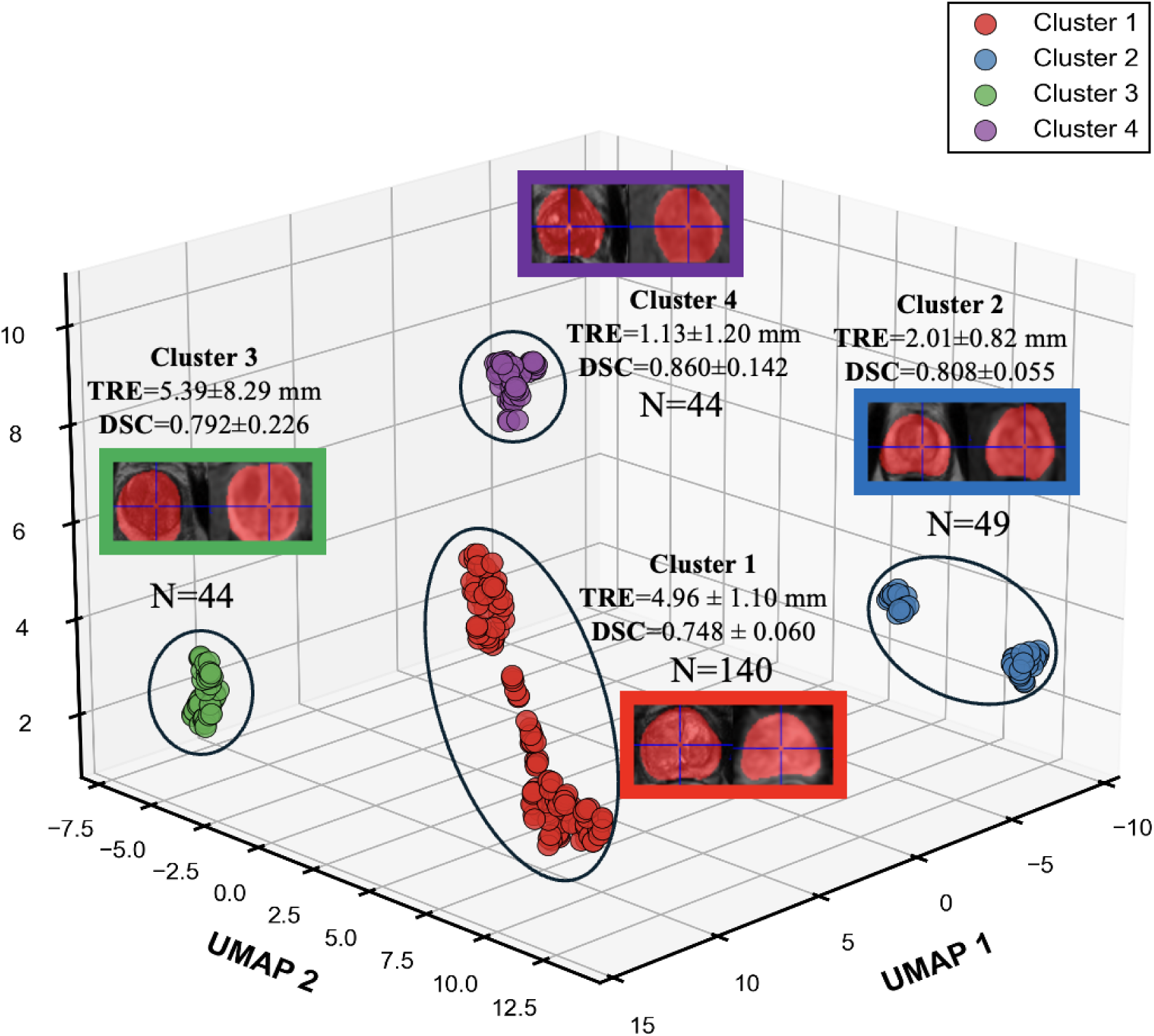
UMAP visualization of MRQy-derived image quality clusters with corresponding registration performance metrics. The scatter plot shows the three-dimensional UMAP embedding of bpMRI scans (n=277) colored by four distinct quality clusters identified through unsupervised clustering of MRQy metrics. Each cluster is annotated with representative T2-weighted (fixed) and ADC image (moved) pairs along with their corresponding Target Registration Error (TRE) and Dice Similarity Coefficient (DSC) values.

## Results

### PI-QUAL based assessment of quality of MRI scans

Among the 568 bpMRI scans from seven cohorts, 277 were classified as high-quality according to PI-QUAL v2 criteria. The distribution of high PI-QUAL group across institutions is summarized in Table 1. Scans were acquired on MRI systems from GE, Siemens, and Philips at 3T with pelvic phased array coils, except for 9 cases performed with an additional endorectal coil (Table S1).

### Experiment 1. MRQy detects site specific batch effects in PI-QUAL defined defined high-quality scans

For the high PI-QUAL bpMRI subset, unsupervised clustering yielded four clusters (C_1_–C_4_) that closely corresponded to site of acquisition with an overall overlap accuracy of 89.9% (Figure S1). The institutional composition of each cluster (C_1_–C_4_) is shown in Table 1.

### Experiment 2. Impact on the generalizability of prostate gland segmentation due to batch effects

#### Single-Cluster Training

The performance of M₁, when trained on individual clusters and evaluated across all other clusters, revealed substantial variability in segmentation accuracy. The model trained on C₁ (n=140) demonstrated the highest training performance (training DSC=0.91±0.04) and achieved the best cross-cluster generalization with a mean DSC of 0.85±0.02 (range: 0.82-0.89) when evaluated on other clusters. However, statistical analysis revealed highly significant performance variability across evaluation clusters for all metrics (p<0.001), indicating the presence of batch effects despite the good absolute performance. The model achieved balanced precision of 0.85±0.04 and recall of 0.86±0.05, with a volume ratio of 1.03±0.10. The IoU of 0.74±0.04 confirmed robust spatial overlap, while the HD95 of 7.47±0.03 mm and ASSD of 2.18±0.53 mm demonstrated boundary delineation accuracy. The model trained on C₂ (n=49) (training DICE=0.90±0.02) showed dramatically poor cross-cluster generalization, achieving a mean DSC of only 0.41±0.02 (p=0.272) when evaluated on other clusters. The Kruskal-Wallis test revealed no significant difference in DSC performance across different evaluation clusters, suggesting consistent but uniformly poor generalization. The model exhibited extremely high precision of 0.93±0.01 (p=0.978) but severely compromised recall of only 0.29±0.01 (p=0.308), indicating a tendency to under-segment with high specificity. This imbalance resulted in a critically low volume ratio of 0.32±0.02 (p=0.407), demonstrating systematic underestimation of prostate volumes by approximately 68 %. The IoU of 0.28±0.01 (p=0.270) was the lowest among all single-cluster models, reflecting poor spatial overlap. The HD95 of 18.77±0.86 mm (p=0.374) and ASSD of 8.62±0.48 mm (p=0.579) were dramatically elevated, indicating severe boundary detection failures and confirming catastrophic generalization breakdown. The lack of statistical significance across most metrics for C₂ suggests uniformly poor performance regardless of the evaluation cluster. The model trained on C₃ (n=44) (training DICE=0.87±0.11) demonstrated moderate cross-cluster generalization with a mean DSC of 0.71±0.01 (p=0.004). Statistical testing revealed significant differences in DSC (p=0.004), precision (p<0.001), IoU (p=0.004), volume ratio (p=0.011), and ASSD (p=0.002) across evaluation clusters, while recall showed no significant difference (p=0.593), indicating variable performance. The volume ratio of 0.82±0.05 (p=0.011) suggested a tendency toward slight underestimation. The HD95 of 15.25±2.83 mm (p<0.001) and ASSD of 4.30±0.33 mm (p=0.002) indicated moderate boundary accuracy. The model trained on C₄ (n=44) (training DICE=0.82±0.06) showed moderate cross-cluster generalization with a mean DSC of 0.66±0.06 (p<0.001) and the highest variability in performance (standard deviation=0.06). Significant differences were detected across most metrics: DSC (p<0.001), precision (p=0.030), recall (p<0.001), IoU (p<0.001), volume ratio (p<0.001), HD95 (p<0.001), and ASSD (p=0.002). The model achieved precision of 0.82±0.02 but lower recall of 0.60±0.07, indicating a conservative segmentation approach. The volume ratio of 0.76±0.09 demonstrated consistent underestimation across evaluation clusters. The IoU of 0.51±0.06 was the second lowest among single-cluster models. The HD95 of 12.04±1.99 mm and ASSD of 4.88±1.27 mm indicated moderate boundary detection performance (Table 2, Figure S2).

**Table 2.** Cross-cluster segmentation performance metrics for prostate MRI analysis. For each metric, values are presented in three rows: (1) mean ± standard deviation across all samples within the testing clusters; (2) range (minimum–maximum); and (3) *P*-values from Kruskal–Wallis tests comparing each cluster’s metric distribution to that of all other testing clusters combined.

| Training Cluster | Dice | Precision | Recall | Hausdorff Distance (95th percentile) | IoU | Volume_ratio | Average Symmetric Surface Distance |
| --- | --- | --- | --- | --- | --- | --- | --- |
| Cluster 1 (n=140)<br>Training<br>DICE=0.91±0.04 | 0.85±0.02<br>(0.82-0.89)<br>p<0.001*** | 0.85±0.04<br>(0.78-0.89)<br>p<0.001*** | 0.86±0.05<br>(0.79-0.91)<br>p<0.001*** | 7.47±0.03<br>(4.47-10.00)<br>p<0.001*** | 0.74±0.04<br>(0.72-0.80)<br>p<0.001*** | 1.03±0.10<br>(0.90-1.16)<br>p<0.001*** | 2.18±0.53<br>(1.63-2.65)<br>p<0.001*** |
| Cluster 2 (n=49)<br>Training<br>DICE=0.90±0.02 | 0.41±0.02<br>(0.39-0.44)<br>p=0.272 | 0.93±0.01<br>(0.91-0.95)<br>p=0.978 | 0.29±0.01<br>(0.27-0.32)<br>p=0.308 | 18.77±0.86<br>(17.90-19.62)<br>p=0.374 | 0.28±0.01<br>(0.26-0.30)<br>p=0.270 | 0.32±0.02<br>(0.30-0.34)<br>p=0.407 | 8.62±0.48<br>(8.22-9.17)<br>p=0.579 |
| Cluster 3 (n=44)<br>Training<br>DICE=0.87±0.11 | 0.71±0.01<br>(0.69-0.74)<br>p=0.004** | 0.83±0.05<br>(0.76-0.89)<br>p<0.001*** | 0.66±0.01<br>(0.65-0.67)<br>p=0.593 | 15.25±2.83<br>(11.93-18.87)<br>p<0.001*** | 0.57±0.02<br>(0.54-0.61)<br>p=0.004** | 0.82±0.05<br>(0.76-0.90)<br>p=0.011* | 4.30±0.33<br>(3.92-4.74)<br>p=0.002** |
| Cluster 4 (n=44)<br>Training<br>DICE=0.82±0.06 | 0.66±0.06<br>(0.58-0.72)<br>p<0.001*** | 0.82±0.02<br>(0.80-0.85)<br>p=0.030* | 0.60±0.07<br>(0.50-0.67)<br>p<0.001** | 12.04±1.99<br>(10.76-14.43)<br>p<0.001*** | 0.51±0.06<br>(0.43-0.57)<br>p<0.001*** | 0.76±0.09<br>(0.63-0.85)<br>p<0.001*** | 4.88±1.27<br>(4.04-6.44)<br>p=0.002** |
| Multi-cluster (n=60)<br>Training<br>DICE=0.84±0.01 | 0.81±0.01<br>(0.79-0.83)<br>p=0.075 | 0.85±0.04<br>(0.80-0.91)<br>p<0.001*** | 0.81±0.05<br>(0.76-0.89)<br>p<0.001*** | 7.44±2.06<br>(5.38-10.36)<br>p=0.008** | 0.70±0.02<br>(0.67-0.72)<br>p=0.075 | 0.97±0.12<br>(0.85-1.15)<br>p<0.001*** | 2.52±0.34<br>(2.07-2.86)<br>p=0.012* |

Significance: \*\*\* p<0.001, \*\* p<0.01, \*p<0.05

#### Balanced Multi-Cluster Training

In contrast to single-cluster models, the balanced multi-cluster model (M₁ trained on n=60 samples selected from all four clusters, achieving training DICE=0.84±0.01) demonstrated superior and more consistent cross-cluster generalization. Statistical analysis revealed no significant differences in the primary segmentation metrics when evaluated across different clusters: DSC (p=0.075) and IoU (p=0.075), indicating robust transferability across diverse prostate MRI phenotypes. The model achieved a mean DSC of 0.81±0.01 (range: 0.79-0.83). The multi-cluster model maintained high and balanced performance across all metrics, with precision of 0.85±0.04 (0.80-0.91, p<0.001) and recall of 0.81±0.05 (0.76-0.89, p<0.001). While precision and recall showed statistically significant differences across evaluation clusters (both p<0.001), their absolute values remained consistently high and well-balanced, avoiding the severe precision-recall trade-offs observed in single-cluster models. The IoU of 0.70±0.02 (0.67-0.72, p=0.075) confirmed robust spatial overlap between predictions and ground truth. The HD95 of 7.44±2.06 mm (5.38-10.36 mm, p=0.008) was comparable to the best-performing single-cluster model, while the ASSD of 2.52±0.34 mm (2.07-2.86 mm, p=0.012) was the second best overall, demonstrating consistent boundary detection accuracy across diverse acquisition protocols. The multi-cluster model achieved a volume ratio of 0.97±0.12 (0.85-1.15, p<0.001), approaching the ideal value of 1.0 and demonstrating near-optimal volumetric accuracy. Although volume ratio showed significant variability across clusters (p<0.001), the mean value and range indicated substantially improved calibration compared to single-cluster models, which exhibited systematic biases toward either minimal overestimation (C₁: 1.03) or substantial underestimation (C₂: 0.32, C₃: 0.82, C₄: 0.76) (Table 2).

To assess whether the multi-cluster model achieved comparable segmentation performance to the best single-cluster model despite using fewer training samples, we conducted equivalence testing using the TOST procedure with a clinically relevant margin of ±0.10 DSC (24). The multi-cluster model showed slightly lower absolute DSC compared to the single-cluster C₁ model (difference: – 0.04). However, the TOST test confirmed statistical equivalence (p<0.001), indicating that this difference lies entirely within clinically acceptable bounds. Having established equivalent performance, we then quantified training efficiency by computing the performance-per-sample ratio using bootstrap resampling. The multi-cluster model achieved 2.23-fold greater training efficiency (95% CI: 2.18-2.29) compared to the C₁ model. These findings demonstrated that the multi-cluster approach achieves equivalent segmentation accuracy while requiring 2.3-fold fewer training samples and eliminating significant batch effects (p=0.075 vs p<0.001) (Table 3).

**Table 3.** Training Efficiency Comparison: Single-Cluster vs Multi-Cluster Approach.

| Parameter | Cluster 1 (n=140) | Multi-cluster (n=60) | Interpretation |
| --- | --- | --- | --- |
| DSC | 0.85±0.02 | 0.81±0.01 | Equivalent (TOST p<0.001) |
| HD95 (mm) | 7.47±0.03 | 7.44±2.06 | Comparable |
| IoU | 0.74±0.04 | 0.70±0.02 | Comparable |
| Generalizability | p<0.001*** | p=0.075 | No batch effect |
| Efficiency ratio | 1.00 (reference) | 2.23 [2.18-2.29] | Efficient |

Abbreviations: DSC, Dice similarity coefficient; HD95, Hausdorff distance 95th percentile; IoU, intersection over union; TOST, two one-sided tests. Cross-cluster variability assessed using Kruskal-Wallis test. DSC equivalence confirmed using TOST procedure with ±0.10 margin. \*\*\*p<0.001.

### Experiment 3: Evaluating Generalizability of Registration Algorithm Due to Batch Effects

M₂ was successfully applied to align ADC maps with their corresponding T2w images across all 277 high PI-QUAL bpMRI scans. Registration performance varied significantly between the different MRQy-identified clusters, with the Kruskal-Wallis test revealing statistically significant differences in both DSC (p=0.047) and TRE (p<0.001) metrics. C₄ demonstrated the highest registration accuracy with a median TRE of 0.86 mm (Inter Quartile Range (IQR)=0.29-1.46 mm). C₂ showed strong performance with DSC of 0.808±0.055 and median TRE of 1.83 mm (IQR: 1.56-2.36 mm). Moderate performance was observed in C₃ with DSC of 0.792±0.226 and median TRE of 1.54 mm (IQR: 0.59-4.20 mm). C₁ demonstrated the lowest registration DSC of 0.748±0.060 with median TRE of 5.47 mm (IQR: 4.59-5.66 mm). Despite these variations, three of four clusters (C_2_, C_3_, C_4_) achieved clinically acceptable registration accuracy with median TRE<2.5 mm and DSC>0.75, while C_1_ demonstrated elevated median TRE of 5.47 mm (IQR: 4.59-5.66) (26).( Table 4, Figure S3).

**Table 4.** Comparison of Registration Performance Across Clusters Using TRE and DSC Metrics.

| Metrics | Subjects | TRE Median [IQR] (mm) | *P* | DSC (Mean±Std) | *P* |
| --- | --- | --- | --- | --- | --- |
| TRE | Cluster 1 | 5.47 [4.59-5.66] | <0.001 | 0.748 ± 0.060 | 0.047 |
|  | Cluster 2 | 1.83 [1.56-2.36] |  | 0.808 ± 0.055 |  |
|  | Cluster 3 | 1.54 [0.59-4.20] |  | 0.792 ± 0.226 |  |
|  | Cluster 4 | 0.86 [0.29-1.46] |  | 0.860 ± 0.142 |  |

*Significance level was set at $P < 0.05$ for Kruskal-Wallis test*

## Discussion

This study demonstrated that acquisition-related batch effects can significantly impair the performance and generalizability of AI algorithms in prostate MRI analysis, even when scans meet standardized high-quality criteria. Using quantitative metrics from MRQy and unsupervised clustering, we identified site-specific technical patterns not captured by PI-QUAL v2, uncovering hidden sources of variability within visually homogeneous datasets. Through two foundational tasks in prostate MRI analysis, gland segmentation and multi-sequence deformable coregistration, we showed that acquisition variability creates systematic signatures that AI models unintentionally learn, reducing their cross-cluster robustness. Importantly, we found that strategic diversity-aware training resolved batch effects for supervised models. Our balanced multi-cluster approach successfully eliminated batch effects and achieved 2.23-fold greater training efficiency compared to larger single-cluster datasets. This demonstrated that how diversity is incorporated during training matters more than data quantity alone.

For clinical deployment, addressing acquisition-related variability is essential. Our findings revealed that batch effects persist despite standardized protocols and significantly impair model performance across clusters. Prior studies have reported similar challenges. Rouviere et al. showed that scanner type influences prostate segmentation accuracy, and Kushol et al. demonstrated that neuroimaging classification performance declines across vendors despite harmonization methods such as ComBat (14, 15). However, these studies primarily addressed segmentation or harmonization approaches without accounting for image quality, whereas our work systematically evaluates generalizability in a PI-QUAL–curated, multi-institutional prostate MRI cohort. By integrating both visual and quantitative metrics, we demonstrated that acquisition-related variability consistently affects two fundamental tasks in prostate MRI, even within visually homogeneous, guideline-compliant datasets. For segmentation, single-cluster models produced highly variable cross-cluster results with severe under-segmentation. Even the largest single-cluster model exhibited significant batch effects across all metrics, indicating that increasing training data from a single technical environment is insufficient to overcome acquisition-related variability. This finding appears to contrast with Netzer et al. (27), who reported that expanding training data from 171 to 806 cases improved prostate cancer detection, while institutional and scanner diversity provided no additional benefit. However, this discrepancy reflects a critical distinction. Netzer et al. (27) examined institutional boundaries, whereas our study isolated technical diversity within PI-QUAL-defined high-quality scans using quantitative MRQy features. Their findings suggest that institutional labels do not capture the acquisition-level variations our clustering revealed. Despite using 2.3-fold fewer training samples, our balanced multi-cluster model achieved superior generalizability while eliminating significant batch effects and maintaining equivalent performance. These results demonstrate that technical diversity outweighs data quantity for robust AI generalization. To evaluate whether batch effects extend beyond segmentation, we assessed deformable registration for aligning ADC maps to T2-weighted images. Notably, unlike the segmentation experiments where we compared different training strategies, the registration model was trained on all high-quality image pairs across all four clusters, providing comprehensive exposure to technical variability. Despite this exhaustive training, registration demonstrated substantial cluster-dependent performance differences, confirming that batch effects persist across multiple foundational tasks. This divergence in outcomes between segmentation and registration highlights the critical role of the learning paradigm. Diversity-aware training with ground truth supervision successfully eliminated batch effects for segmentation, as the supervised loss signal enabled the model to learn cluster-invariant features for anatomical boundary detection. In contrast, unsupervised registration faced fundamental limitations that prevented it from overcoming batch effects despite exposure to all cluster types during training. First, unsupervised registration lacks ground truth deformation fields to guide learning. Instead, the network optimizes similarity metrics (e.g., normalized mutual information) as proxy objectives, learning transformations that maximize image similarity rather than anatomical correspondence. When these metrics are differentially reliable across clusters; the model has no mechanism to correct cluster-specific failures. Second, the network learns a single universal transformation function that must simultaneously accommodate all cluster types. This universal function compromises across different acquisition characteristics and cannot dynamically adapt to cluster-specific requirements. Acquisitions with large slice-thickness disparities may require different deformation strategies than those with endorectal coil distortions or b-value variations, but the unsupervised training objective provides no signal to learn these distinctions. These findings suggest that batch effect mitigation strategies must be tailored to the learning paradigm. Diversity-aware sampling successfully addresses batch effects in supervised tasks where ground truth labels provide explicit feedback. Unsupervised tasks lacking anatomical guidance may require alternative approaches, such as cluster-specific models.

Our findings have important implications for the clinical implementation of AI in prostate MRI. Cluster-guided sampling provides a practical strategy for assembling technically diverse training cohorts without requiring prohibitively large datasets. In our study, the multi-cluster model achieved equivalent segmentation performance to the single-cluster model while eliminating significant batch effects and demonstrating a 2.23-fold increase in training efficiency, challenging the assumption that larger datasets inherently yield better performance (27). In practice, targeting approximately 15 to 20 cases per technical cluster appears sufficient to capture dominant acquisition variability, offering clearer guidance for dataset curation than simply maximizing total sample size. For AI developers, integrating quantitative tools such as MRQy into data pipelines enables prospective identification of technical clusters and balanced sampling during model construction. Deploying institutions may benefit from lightweight pre-deployment validation on a small set of representative local scans to detect systematic biases when acquisition protocols differ from those in the training data. Clinically, incorporating image-quality metrics into radiology workflows can support real-time recognition of out-of-distribution scans, helping to provide confidence indicators for AI outputs and informing decisions about when manual review is warranted. These recommendations are aligned with widely discussed perspectives from regulatory bodies. The United States Food and Drug Administration (FDA) highlight the importance of demonstrating generalizable performance across the intended patient population and across relevant imaging conditions. Public evaluations of FDA-cleared AI systems indicate that many submissions still include limited external validation or subgroup reporting (28, 29). In Europe, the regulatory frameworks emphasize the need to show that AI systems perform reliably in the target clinical context, and recent European Union policy initiatives such as the EU AI Act introduce requirements for transparency, bias mitigation, and data governance for high-risk medical AI systems (30). By combining quantitative clustering, equivalence testing, and efficiency metrics, our framework provides transparent and reproducible benchmarks that can support both regulatory submissions and post-deployment monitoring.

Several limitations of this study should be acknowledged. First, our analysis emphasized technical metrics for segmentation and registration rather than diagnostic accuracy or clinically relevant endpoints such as identifying clinically significant prostate cancer. This reflects both the limited availability of comprehensive histopathologic data and our primary objective of determining whether hidden acquisition-related variability affects AI model performance, a question best addressed through foundational technical tasks. Regarding generalizability to diagnostic tasks, segmentation and registration rely on similar low-level image features affected by acquisition parameters, and clinical AI systems for prostate cancer detection typically incorporate both tasks as essential preprocessing steps (21). Batch effects impairing these foundational tasks would inevitably propagate to downstream diagnostic predictions. Additionally, the technical factors driving batch effects in our study, including variations in signal-to-noise ratio, spatial resolution, and intensity distributions, are the same image characteristics that influence lesion conspicuity and ADC quantification essential for cancer detection. However, important differences may exist between proxy and diagnostic tasks. Lesion detection models focusing on focal abnormalities might show different sensitivity to specific batch effects compared to segmentation tasks requiring precise boundary delineation across the entire gland. For example, subtle variations in ADC values due to b-value selection could disproportionately affect cancer detection performance, since ADC measurements are important for lesion characterization, while having minimal impact on anatomical segmentation. Conversely, geometric distortions that severely impair registration accuracy might minimally affect lesion classification if the lesion remains within the field of view. Furthermore, cancer detection involves higher-level texture and morphological features that may exhibit different robustness to acquisition variability compared to the boundary-sensitive features used for segmentation. Future work should explicitly evaluate batch effects on histopathological confirmed cancer detection endpoints to quantify diagnostic impact and determine whether diversity-aware training strategies provide similar benefits for diagnostic tasks. Second, our segmentation models achieved Dice scores ranging from 0.82 to 0.91 for the best performing approaches, which, while clinically acceptable, are below state of the art benchmarks reported in dedicated segmentation challenges such as PROMISE12 and PROSTATEx (31, 32). This relatively lower performance may partially reflect the technical heterogeneity inherent in our multi-institutional dataset compared to more controlled challenge datasets and may have amplified the observed batch effects. However, this limitation does not invalidate our central finding that technical diversity in training data reduces cross-cluster performance variability, a pattern that would likely persist regardless of absolute performance levels. Third, regarding the relationship between technical clusters and institutional sites, our MRQy-defined clusters showed 89.9% concordance with acquisition sites. We acknowledge that we did not directly compare technical cluster-balanced training with site-balanced training, which represents a limitation of our study. While this high concordance raises the question of whether technical clustering adds value over simple site-based stratification, we emphasize that technical clustering captures quantitative imaging characteristics rather than simply capturing site labels. This distinction is meaningful for several reasons: (1) technical features can generalize to new institutions by applying the same quantitative framework, enabling prospective deployment without requiring site-specific metadata; (2) technical clustering can detect within-site temporal variations from scanner upgrades or protocol changes that site-based stratification would miss; (3) quantitative diversity metrics provide objective, transparent documentation of dataset heterogeneity suitable for regulatory submissions; and (4) technical clustering identifies underlying acquisition mechanisms (e.g., SNR differences, intensity distributions) rather than arbitrary institutional labels, providing insights into which specific technical factors drive batch effects. Nevertheless, the central contribution of our work is to demonstrate that strategic diversity-aware sampling improves generalizability compared with naive pooling of all available data, regardless of whether diversity is defined by technical clusters or by institutional site. Future work should directly compare site-based and cluster-based stratification strategies to quantify the incremental value of quantitative technical characterization. Fourth, the study included a modest number of institutions and scanner types, which may not fully capture acquisition variability in global clinical practice. Additionally, uneven distribution of cases across clusters may have reduced statistical power. To address this concern, we note that our balanced multi-cluster model outperformed the single-cluster model despite training on fewer cases, and the efficiency analysis with bootstrap confidence intervals provides robust statistical evidence independent of absolute sample sizes. Fifth, we did not evaluate alternative strategies for mitigating batch effects, such as adversarial domain adaptation, style transfer, or statistical harmonization methods like ComBat (33). These approaches offer complementary benefits and limitations. ComBat and related techniques harmonize images by adjusting intensities to remove site-specific variations but risk removing biologically relevant variations alongside technical artifacts (34). Domain adaptation methods require labeled data from target domains and increase computational complexity (33). Style transfer approaches face challenges in defining appropriate reference styles and validating transformed images (33). Our approach offers key advantages preserving original unmodified images ensures interpretability; the method requires only unsupervised clustering during dataset curation with no modifications to standard training algorithms; and quantitative diversity metrics provide transparent documentation of dataset representativeness valuable for regulatory submissions. Finally, our analysis was limited to bpMRI and did not include dynamic contrast-enhanced (DCE) imaging. While PIRADS and PI-QUAL criteria are increasingly applied to bpMRI, the absence of DCE may limit generalizability. Future work will seek to examine whether similar batch effects persist in mpMRI datasets and will seek to evaluate their impact on DCE-derived parameters, which are known to be variable and may be differentially affected (35).

Despite the aforementioned limitations, this study provides clear evidence that acquisition-related batch effects significantly impair the generalizability of AI models for prostate MRI analysis, even when scans meet high visual quality standards. Using quantitative image quality metrics and unsupervised clustering, we identified hidden technical variability within PI-QUAL compliant datasets that conventional quality assessment fails to capture, with both segmentation and registration tasks demonstrating substantial performance degradation across technical clusters. Importantly, strategic diversity-aware training effectively mitigated batch effects for supervised learning, with our balanced multi-cluster approach eliminating significant batch effects while achieving superior training efficiency compared to larger single-cluster datasets. This challenges the conventional assumption that simply increasing dataset size improves performance and suggests that targeting technical diversity may be more effective than maximizing sample size. As AI systems transition from research to clinical deployment, robust multi-site training strategies, prospective quality monitoring, and transparent reporting of technical diversity will be critical to ensuring equitable, accurate, and trustworthy performance across diverse imaging environments.

## Data Availability Statement

The code for MRQy-based clustering, segmentation and registration model trainings, and statistical analysis are publicly available at https://github.com/Emory-Empathetic-AI-for-Health-Inst/Multimodal-Multi-scale-Framework-for-Ethical-AI-Model-Development. The in-house imaging data cannot be shared publicly due to institutional privacy regulations and patient consent restrictions; however, de-identified data may be made available upon reasonable request and institutional approval.

## Data Sharing Statement

The data underlying this article are available from the corresponding author upon reasonable request. Publicly available data from the PROSTATEx challenge were also used in this study.

## Author contributions

Guarantor of integrity of entire study, S.A., A.Ma.; study concepts/study design, S.A., A.S., A.Ma.; data acquisition, S.A., Z.Z., M.G., F.H., P.S., A.R.R., S.V., E.G.K., M.S., M.H., R.S.; data analysis/interpretation, S.A., A.S., A.Ma., A.Mi., Z.Z.; manuscript drafting, S.A., A.S., A.Ma.; manuscript revision for important intellectual content, all authors; approval of final version of submitted manuscript, all authors; agrees to ensure any questions related to the work are appropriately resolved, all authors; literature research, S.A., A.S., A.Mi., Z.Z., F.H., P.S.; statistical analysis, S.A..; manuscript editing, all authors.

## Title of the Manuscript

Evaluating the Interplay and Impact of Quality and Technical Variations on Performance of AI Models in Multi-Institutional Prostate MRI Datasets

## Abbreviations

AI: Artificial intelligence
bpMRI: Biparametric prostate MRI
DSC: Dice similarity coefficient
IoU: Intersection over union
TRE: Target registration error
PI-QUAL: Prostate Imaging Quality

## References

1. Bergengren O, Pekala KR, Matsoukas K, Fainberg J, Mungovan SF, Bratt O, Bray F, Brawley O, Luckenbaugh AN, Mucci L, Morgan TM, Carlsson SV. 2022 Update on Prostate Cancer Epidemiology and Risk Factors-A Systematic Review. Eur Urol 2023;84(2):191-206. doi: 10.1016/j.eururo.2023.04.021

2. Pinsky PF, Parnes H. Screening for Prostate Cancer. N Engl J Med 2023;388(15):1405–1414. doi: 10.1056/NEJMcp2209151

3. Schaeffer EM, Srinivas S, Adra N, An Y, Barocas D, Bitting R, Bryce A, Chapin B, Cheng HH, D’Amico AV, Desai N, Dorff T, Eastham JA, Farrington TA, Gao X, Gupta S, Guzzo T, Ippolito JE, Kuettel MR, Lang JM, Lotan T, McKay RR, Morgan T, Netto G, Pow-Sang JM, Reiter R, Roach M, Robin T, Rosenfeld S, Shabsigh A, Spratt D, Teply BA, Tward J, Valicenti R, Wong JK, Shead DA, Snedeker J, Freedman-Cass DA. Prostate Cancer, Version 4.2023, NCCN Clinical Practice Guidelines in Oncology. J Natl Compr Canc Netw 2023;21(10):1067–1096. doi: 10.6004/jnccn.2023.0050

4. Weinreb JC, Barentsz JO, Choyke PL, Cornud F, Haider MA, Macura KJ, Margolis D, Schnall MD, Shtern F, Tempany CM, Thoeny HC, Verma S. PI-RADS Prostate Imaging – Reporting and Data System: 2015, Version 2. Eur Urol 2016;69(1):16–40. doi: 10.1016/j.eururo.2015.08.052

5. Turkbey B, Rosenkrantz AB, Haider MA, Padhani AR, Villeirs G, Macura KJ, Tempany CM, Choyke PL, Cornud F, Margolis DJ, Thoeny HC, Verma S, Barentsz J, Weinreb JC. Prostate Imaging Reporting and Data System Version 2.1: 2019 Update of Prostate Imaging Reporting and Data System Version 2. Eur Urol 2019;76(3):340–351. doi: 10.1016/j.eururo.2019.02.033

6. Oerther B, Nedelcu A, Engel H, Schmucker C, Schwarzer G, Brugger T, Schoots IG, Eisenblaetter M, Sigle A, Gratzke C, Bamberg F, Benndorf M. Update on PI-RADS Version 2.1 Diagnostic Performance Benchmarks for Prostate MRI: Systematic Review and Meta-Analysis. Radiology 2024;312(2):e233337. doi: 10.1148/radiol.233337

7. Cheng Y, Zhang L, Wu X, Zou Y, Niu Y, Wang L. Impact of prostate MRI image quality on diagnostic performance for clinically significant prostate cancer (csPCa). Abdom Radiol (NY) 2024;49(11):4113–4124. doi: 10.1007/s00261-024-04458-7

8. de Rooij M, Allen C, Twilt JJ, Thijssen LCP, Asbach P, Barrett T, Brembilla G, Emberton M, Gupta RT, Haider MA, Kasivisvanathan V, Logager V, Moore CM, Padhani AR, Panebianco V, Puech P, Purysko AS, Renard-Penna R, Richenberg J, Salomon G, Sanguedolce F, Schoots IG, Thony HC, Turkbey B, Villeirs G, Walz J, Barentsz J, Giganti F. PI-QUAL version 2: an update of a standardised scoring system for the assessment of image quality of prostate MRI. Eur Radiol 2024. doi: 10.1007/s00330-024-10795-4

9. Saha A, Bosma JS, Twilt JJ, van Ginneken B, Bjartell A, Padhani AR, Bonekamp D, Villeirs G, Salomon G, Giannarini G, Kalpathy-Cramer J, Barentsz J, Maier-Hein KH, Rusu M, Rouviere O, van den Bergh R, Panebianco V, Kasivisvanathan V, Obuchowski NA, Yakar D, Elschot M, Veltman J, Futterer JJ, de Rooij M, Huisman H, consortium P-C. Artificial intelligence and radiologists in prostate cancer detection on MRI (PI-CAI): an international, paired, non-inferiority, confirmatory study. Lancet Oncol 2024;25(7):879–887. doi: 10.1016/S1470-2045(24)00220-1

10. Schmitt M, Maron RC, Hekler A, Stenzinger A, Hauschild A, Weichenthal M, Tiemann M, Krahl D, Kutzner H, Utikal JS, Haferkamp S, Kather JN, Klauschen F, Krieghoff-Henning E, Fröhling S, von Kalle C, Brinker TJ. Hidden Variables in Deep Learning Digital Pathology and Their Potential to Cause Batch Effects: Prediction Model Study. J Med Internet Res 2021;23(2):e23436. doi: 10.2196/23436

11. Wu Y, Xu X, Cheng Y, Zhang X, Liu F, Li Z, Hu L, Madabhushi A, Gao P, Liu Z, Lu C. BEEx Is an Open-Source Tool That Evaluates Batch Effects in Medical Images to Enable Multicenter Studies. Cancer Res 2025;85(2):218–230. doi: 10.1158/0008-5472.CAN-23-3846

12. Poltronieri TS, de Paula NS, Chaves GV. Assessing skeletal muscle radiodensity by computed tomography: An integrative review of the applied methodologies. Clin Physiol Funct Imaging 2020;40(4):207–223. doi: 10.1111/cpf.12629

13. Hosny A, Parmar C, Quackenbush J, Schwartz LH, Aerts H. Artificial intelligence in radiology. Nat Rev Cancer 2018;18(8):500–510. doi: 10.1038/s41568-018-0016-5

14. Rouvière O, Moldovan PC, Vlachomitrou A, Gouttard S, Riche B, Groth A, Rabotnikov M, Ruffion A, Colombel M, Crouzet S, Weese J, Rabilloud M. Combined model-based and deep learning-based automated 3D zonal segmentation of the prostate on T2-weighted MR images: clinical evaluation. Eur Radiol 2022;32(5):3248–3259. doi: 10.1007/s00330-021-08408-5

15. Kushol R, Parnianpour P, Wilman AH, Kalra S, Yang YH. Effects of MRI scanner manufacturers in classification tasks with deep learning models. Sci Rep 2023;13(1):16791. doi: 10.1038/s41598-023-43715-5

16. Rajpurkar P, Lungren MP. The Current and Future State of AI Interpretation of Medical Images. N Engl J Med 2023;388(21):1981–1990. doi: 10.1056/NEJMra2301725

17. Sadri AR, Janowczyk A, Zhou R, Verma R, Beig N, Antunes J, Madabhushi A, Tiwari P, Viswanath SE. Technical Note: MRQy – An open-source tool for quality control of MR imaging data. Med Phys 2020;47(12):6029–6038. doi: 10.1002/mp.14593

18. Twilt JJ, Saha A, Bosma JS, van Ginneken B, Bjartell A, Padhani AR, Bonekamp D, Villeirs G, Salomon G, Giannarini G, Kalpathy-Cramer J, Barentsz J, Maier-Hein KH, Rusu M, Rouvière O, van den Bergh R, Panebianco V, Kasivisvanathan V, Obuchowski NA, Yakar D, Elschot M, Veltman J, Fütterer JJ, Huisman H, de Rooij M. Evaluating Biparametric Versus Multiparametric Magnetic Resonance Imaging for Diagnosing Clinically Significant Prostate Cancer: An International, Paired, Noninferiority, Confirmatory Observer Study. Eur Urol 2025;87(2):240–250. doi: 10.1016/j.eururo.2024.09.035

19. Hamzaoui D, Montagne S, Granger B, Allera A, Ezziane M, Luzurier A, Quint R, Kalai M, Ayache N, Delingette H, Renard-Penna R. Prostate volume prediction on MRI: tools, accuracy and variability. Eur Radiol 2022;32(7):4931–4941. doi: 10.1007/s00330-022-08554-4

20. Wang S, Burtt K, Turkbey B, Choyke P, Summers RM. Computer aided-diagnosis of prostate cancer on multiparametric MRI: a technical review of current research. Biomed Res Int 2014;2014:789561. doi: 10.1155/2014/789561

21. Kovacs B, Netzer N, Baumgartner M, Schrader A, Isensee F, Weißer C, Wolf I, Görtz M, Jaeger PF, Schütz V, Floca R, Gnirs R, Stenzinger A, Hohenfellner M, Schlemmer HP, Bonekamp D, Maier-Hein KH. Addressing image misalignments in multi-parametric prostate MRI for enhanced computer-aided diagnosis of prostate cancer. Sci Rep 2023;13(1):19805. doi: 10.1038/s41598-023-46747-z

22. Healy J, McInnes L. Uniform manifold approximation and projection. Nature Reviews Methods Primers 2024;4(1):82. doi: 10.1038/s43586-024-00363-x

23. Milletari F, Navab N, Ahmadi SA. V-Net: Fully Convolutional Neural Networks for Volumetric Medical Image Segmentation. Int Conf 3d Vision 2016:565–571. doi: 10.1109/3dv.2016.79

24. Becker AS, Chaitanya K, Schawkat K, Muehlematter UJ, Hötker AM, Konukoglu E, Donati OF. Variability of manual segmentation of the prostate in axial T2-weighted MRI: A multi-reader study. Eur J Radiol 2019;121:108716. doi: 10.1016/j.ejrad.2019.108716

25. Balakrishnan GaZ, Amy and Sabuncu, Mert R. and Guttag, John and Dalca, Adrian V. VoxelMorph: A Learning Framework for Deformable Medical Image Registration. IEEE Transactions on Medical Imaging 2019; p. 1788–1800.

26. Karnik VV, Fenster A, Bax J, Cool DW, Gardi L, Gyacskov I, Romagnoli C, Ward AD. Assessment of image registration accuracy in three-dimensional transrectal ultrasound guided prostate biopsy. Med Phys 2010;37(2):802–813. doi: 10.1118/1.3298010

27. Netzer N, Weißer C, Schelb P, Wang X, Qin X, Görtz M, Schütz V, Radtke JP, Hielscher T, Schwab C, Stenzinger A, Kuder TA, Gnirs R, Hohenfellner M, Schlemmer HP, Maier-Hein KH, Bonekamp D. Fully Automatic Deep Learning in Bi-institutional Prostate Magnetic Resonance Imaging: Effects of Cohort Size and Heterogeneity. Invest Radiol 2021;56(12):799–808. doi: 10.1097/rli.0000000000000791

28. Warraich HJ, Tazbaz T, Califf RM. FDA Perspective on the Regulation of Artificial Intelligence in Health Care and Biomedicine. Jama 2025;333(3):241–247. doi: 10.1001/jama.2024.21451

29. Windecker D, Baj G, Shiri I, Kazaj PM, Kaesmacher J, Gräni C, Siontis GCM. Generalizability of FDA-Approved AI-Enabled Medical Devices for Clinical Use. JAMA Netw Open 2025;8(4):e258052. doi: 10.1001/jamanetworkopen.2025.8052

30. Kalodanis K, Feretzakis G, Rizomiliotis P, Verykios VS, Papapavlou C, Skrekas A, Anagnostopoulos D. Evaluating the Impact of the EU AI Act on Medical Device Regulation. Stud Health Technol Inform 2025;323:40–44. doi: 10.3233/shti250045

31. Litjens G, Toth R, van de Ven W, Hoeks C, Kerkstra S, van Ginneken B, Vincent G, Guillard G, Birbeck N, Zhang J, Strand R, Malmberg F, Ou Y, Davatzikos C, Kirschner M, Jung F, Yuan J, Qiu W, Gao Q, Edwards PE, Maan B, van der Heijden F, Ghose S, Mitra J, Dowling J, Barratt D, Huisman H, Madabhushi A. Evaluation of prostate segmentation algorithms for MRI: the PROMISE12 challenge. Med Image Anal 2014;18(2):359–373. doi: 10.1016/j.media.2013.12.002

32. Tian Z, Li X, Zheng Y, Chen Z, Shi Z, Liu L, Fei B. Graph-convolutional-network-based interactive prostate segmentation in MR images. Med Phys 2020;47(9):4164–4176. doi: 10.1002/mp.14327

33. Hu F, Chen AA, Horng H, Bashyam V, Davatzikos C, Alexander-Bloch A, Li M, Shou H, Satterthwaite TD, Yu M, Shinohara RT. Image harmonization: A review of statistical and deep learning methods for removing batch effects and evaluation metrics for effective harmonization. Neuroimage 2023;274:120125. doi: 10.1016/j.neuroimage.2023.120125

34. Tassi E, Bianchi AM, Calesella F, Vai B, Bellani M, Nenadić I, Piras F, Benedetti F, Brambilla P, Maggioni E. Assessment of ComBat Harmonization Performance on Structural Magnetic Resonance Imaging Measurements. Hum Brain Mapp 2024;45(18):e70085. doi: 10.1002/hbm.70085

35. A Multi-Institutional Comparison of Dynamic Contrast-Enhanced Magnetic Resonance Imaging Parameter Calculations. Sci Rep 2017;7(1):11185. doi: 10.1038/s41598-017-11554-w

