## Supplemental Information for "Evaluating the Interplay and Impact of Quality and Technical Variations on Performance of AI Models in Multi-Institutional Prostate MRI Datasets"

**Supplementary Material:**

**Imaging Protocols**

***Site of Acquisition 1***

All scans were acquired on 3T Siemens Skyra scanners using pelvic phased-array coils, without endorectal coils. T2-weighted imaging was performed using a TSE sequence with a TR of 7,500 ms, TE of 101 ms, and a flip angle of 160°. Axial images were acquired with a slice thickness of 3.5 mm and spacing of 4.025 mm, and an in-plane resolution of 0.78 × 0.78 mm (matrix = 320 × 320; FOV ≈ 250 mm). DWI was performed using an SE-EPI sequence with TR of 4,300 ms, TE of 60 ms, and fat suppression. The in-plane resolution was 1.52 × 1.52 mm (matrix = 114 × 114; FOV ≈ 174 mm), with a slice thickness of 3.5 mm and spacing of 4.025 mm. b-values of 0, 800, and 1,500 s/mm² were used.

***Site of Acquisition 2***

All MRI studies were performed on 3T Philips Ingenia and Siemens Skyra scanners with pelvic phased-array surface coils, without endorectal coils.

For Siemens Skyra, T2-weighted imaging was performed using a TSE sequence with a TR of 3,894 ms and TE of 90 ms. The in-plane resolution was 0.56 mm and the slice thickness was 2 mm. DWI was acquired using a single-shot EPI sequence with a TR of 3,656 ms and TE of 64 ms, achieving an in-plane resolution of 0.87 mm and a slice thickness of 3 mm. b-values of 0, 400, 900, 1,000, and 1,500 s/mm² were used.

For Philips Ingenia, T2-weighted imaging was performed using a TSE sequence with a TR of 8,740 ms and TE of 150 ms. The in-plane resolution was 1.25 mm with a slice thickness of 3.5 mm. DWI was performed using a single-shot EPI sequence with a TR of 11,300 ms and TE of 83 ms, achieving an in-plane resolution of 1.5 mm and a slice thickness of 3.5 mm. b-values of 0, 500, 1,500, and 2,000 s/mm² were used.

***Site of Acquisition 3***

All scans were acquired using 3T Siemens Skyra scanners with pelvic phased-array coils, without the use of endorectal coils. T2-weighted imaging was performed using a TSE sequence with a TR of 3,800 ms, TE of 119 ms, and a flip angle of 160°. Images were acquired in the axial plane with a slice thickness of 3 mm, no interslice gap, and an in-plane resolution of 0.98 × 0.98 mm (matrix = 256 × 256; FOV ≈ 250 mm). DWI was performed using a spin echo EPI (SE-EPI) sequence with TR of 4,656 ms and TE of 46 ms. The in-plane resolution was 1.37 × 1.37 mm (matrix = 128 × 128; FOV ≈ 174 mm) with a slice thickness of 3 mm and interslice spacing of 3.3 mm. Parallel imaging and fat suppression were used. b-values of 0, 800, and 1,500 s/mm² were applied.

***Site of Acquisition 4***

All MRI studies were performed using a 3T GE SIGNA Pioneer scanner equipped with a phased-array pelvic coil, without endorectal coils. T2-weighted imaging was acquired using a fast spin echo (FSE) sequence with a TR of 4,996 ms, TE of 168 ms, and a flip angle of 140°. Axial images were acquired with a slice thickness of 3 mm, no interslice gap, and an in-plane resolution of approximately 0.49 × 0.49 mm (matrix = 500 × 512; FOV ≈ 245 mm). A 16-channel anterior and 32-channel posterior coil array was used. DWI was performed using an SE-EPI sequence with TR of 4,461 ms, TE of 59 ms, and a flip angle of 90°. Axial images were acquired with a slice thickness of 3 mm, interslice spacing of 3.3 mm, and an in-plane resolution of approximately 1.36 × 1.36 mm (matrix = 71 × 128; FOV ≈ 174 mm). Partial Fourier imaging (~54.9% undersampling) and an echo train length of 39 were used. b-values of 0, 800, and 1,500 s/mm² were applied, and ADC maps were calculated from three b-values.

***Site of Acquisition 5***

All MRI studies were performed using a 3T Siemens Verio scanner equipped with a 16-channel anterior pelvic coil and an endorectal coil. T2-weighted imaging was performed using a TSE sequence with a TR of 4,000 ms, TE of 120 ms, and a flip angle of 125°. Axial images were acquired with a slice thickness of 3 mm, no interslice gap, and an in-plane resolution of approximately 0.78 × 0.78 mm (matrix = 384 × 384; FOV ≈ 300 mm). DWI was performed using an SE-EPI sequence with monopolar diffusion gradients, TR of 3,795 ms, TE of 96 ms, and a flip angle of 90°. The in-plane resolution was approximately 1.73 × 1.73 mm (matrix = 81 × 116; FOV ≈ 200 mm) with a slice thickness of 3 mm and no interslice gap. Fat suppression, partial Fourier acquisition (75%), and parallel imaging (R = 2) were applied. b-values of 0, 50, 500, 1,000, and 1,500 s/mm² were used, and ADC maps were calculated from b = 0, 50, 500, and 1,000 s/mm².

***Site of Acquisition 6***

All MRI studies were performed using 3T Siemens Skyra scanners equipped with pelvic phased-array surface coils, without endorectal coils. T2-weighted imaging was acquired using TSE sequences with a TR of 4,170 ms and TE of 107 ms. Axial images were acquired with a slice thickness of 3.5 mm, spacing of 4.025 mm, and an in-plane resolution of 0.86 × 0.86 mm. DWI was acquired using SE-EPI sequences with fat suppression and partial Fourier acquisition (~43.9% undersampling), TR of 11,400 ms, and TE of 97 ms. Axial DWI images had a slice thickness of 3.5 mm, spacing of 4.025 mm, and an in-plane resolution of approximately 1.88 × 1.88 mm (matrix = 160 × 102). b-values of 0, 800, 1,000, and 2,000 s/mm² were used, and ADC maps were calculated from b = 0, 800, and 1,000 s/mm².

***Site of Acquisition 7***

All MRI studies were performed on 3T Siemens scanners (MAGNETOM Trio and Skyra) without the use of endorectal coils. T2-weighted images were acquired using a turbo spin echo (TSE) sequence with an in-plane resolution of 0.5 mm and a slice thickness of 3.6 mm. Diffusion-weighted imaging (DWI) was performed using a single-shot echo planar imaging (EPI) sequence, with an in-plane resolution of 2 mm and a slice thickness of 3.6 mm. Diffusion gradients were applied in three directions, and b-values of 50, 400, and 800 s/mm² were used.

A summary of acquisition parameters across sites is provided in the comparative table (Table S1).

**Table S1.** Summary of MRI acquisition parameters across datasets

| Institution | Scanner | Endorectal Coil Used | T2-weighted Imaging | Diffusion-weighted Imaging |
| --- | --- | --- | --- | --- |
| Site of Acquisition 1 | Siemens Skyra | No | TSE, 0.78 mm in-plane, 3.5 mm slice | SE-EPI, 1.52 mm in-plane, 3.5 mm slice, b=0, 800, 1500 |
| Site of Acquisition 2 Part 1 | Siemens Skyra | No | TSE, 0.56–mm in-plane, 2 mm slice | SE-EPI, 0.87 mm in-plane, 3 mm slice, b=0, 400, 900, 1000, 1500 |
| Site of Acquisition 2 Part 2 | Philips Ingenia | No | TSE, 1.25 mm in-plane, 3.5 mm slice | SE-EPI, 1.5 mm in-plane, 3.5 mm slice, b=0, 500, 1500, 2000 |
| Site of Acquisition 3 | Siemens Skyra | No | TSE, 0.98 mm in-plane, 3 mm slice | SE-EPI, 1.37 mm in-plane, 3 mm slice, b=0, 800, 1500 |
| Site of Acquisition 4 | GE SIGNA Pioneer | No | FSE, 0.49–0.98 mm in-plane, 3 mm slice | SE-EPI, 1.36 mm in-plane, 3 mm slice, b=0, 800, 1500 |
| Site of Acquisition 5 | Siemens Verio | Yes | TSE, 0.78 mm in-plane, 3 mm slice | SE-EPI, 1.73 mm in-plane, 3 mm slice, b=0, 50, 500, 1000, 1500 |
| Site of Acquisition 6 | Siemens Skyra | No | TSE, 0.86 mm in-plane, 3.5 mm slice | SE-EPI, 1.88 mm in-plane, 3.5 mm slice, b=0, 800, 1000, 2000 |
| Site of Acquisition 7 | Siemens Trio and Skyra | No | TSE, 0.5 mm in-plane, 3.6 mm slice | SE-EPI, 2 mm in-plane, 3.6 mm slice, b=50, 400, 800 |

**Supplementary Analyses**

This document provides additional analyses and visualizations supporting the main manuscript. Figures illustrate the impact of acquisition-related batch effects on biparametric prostate MRI (bpMRI) and demonstrate how clustering and balanced training improve model robustness.

**Figure S1:** Three-dimensional UMAP projection of 277 PI-QUAL-defined high-quality biparametric prostate MRI scans based on quantitative image quality metrics.

**Figure S2: Comprehensive cross-cluster generalization analysis**

**Figure S3: Registration performance varies significantly across MRQy-defined image quality clusters.**

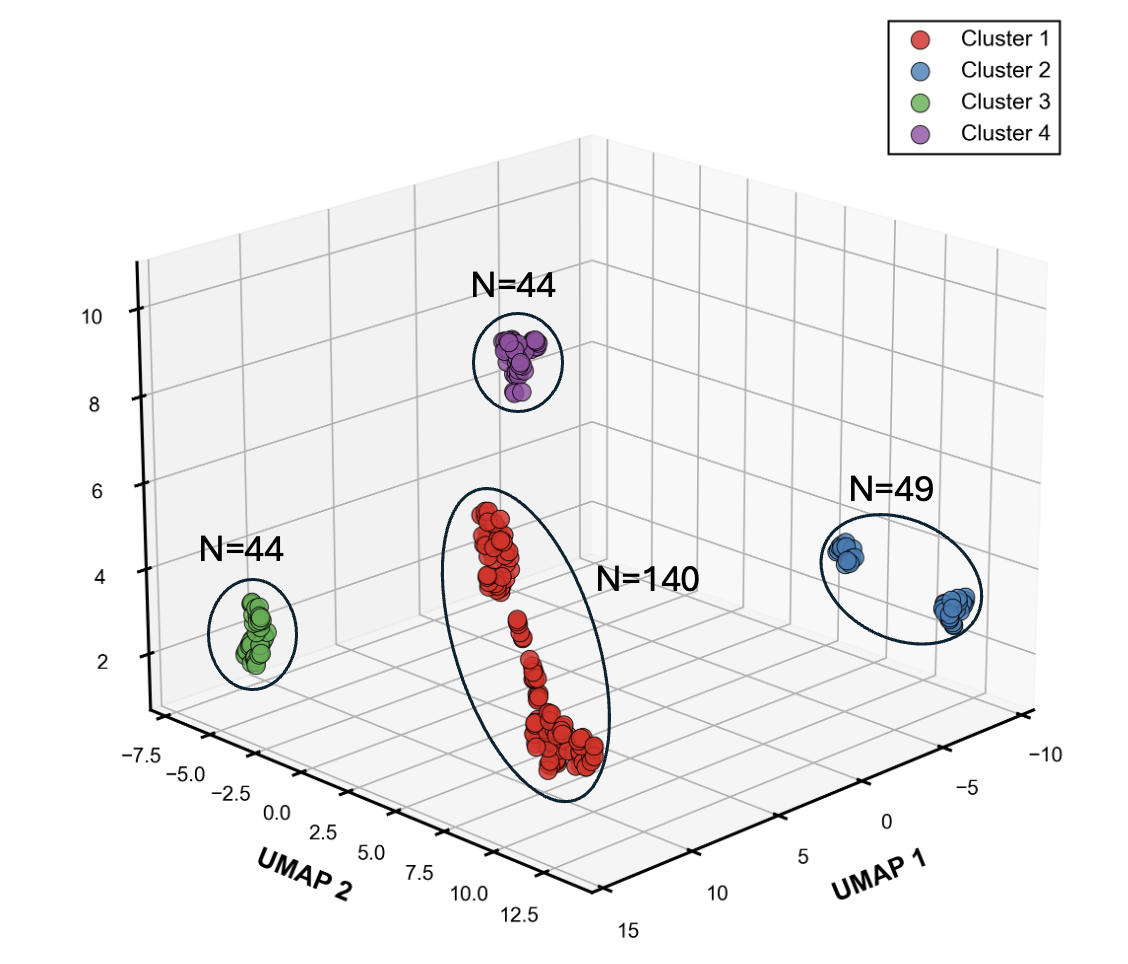
**Figure S1.** Three-dimensional UMAP projection of 277 PI-QUAL-defined high-quality biparametric prostate MRI scans based on quantitative image quality metrics. **UMAP clustering reveals hidden technical heterogeneity within high-quality prostate MRI scans.** K-means clustering identified four distinct technical clusters: Cluster 1 (red, n=140), Cluster 2 (blue, n=49), Cluster 3 (green, n=44), and Cluster 4 (purple, n=44). The spatial separation of clusters demonstrates that acquisition-related batch effects persist despite visual quality homogeneity, with clusters corresponding to site-specific acquisition patterns (89.9% overlap accuracy).

When evaluated by institutions among high-quality bpMRI scans, the cluster overlap accuracy was 92.5% for *Site of Acquisition 1*, 69.0% for *Site of Acquisition 2*, 100% for *Site of Acquisition 3*, 100% for *Site of Acquisition 4*, 100% for *Site of Acquisition 5*, 87.9% for *Site of Acquisition 6*, and 100% for *Site of Acquisition 7*.

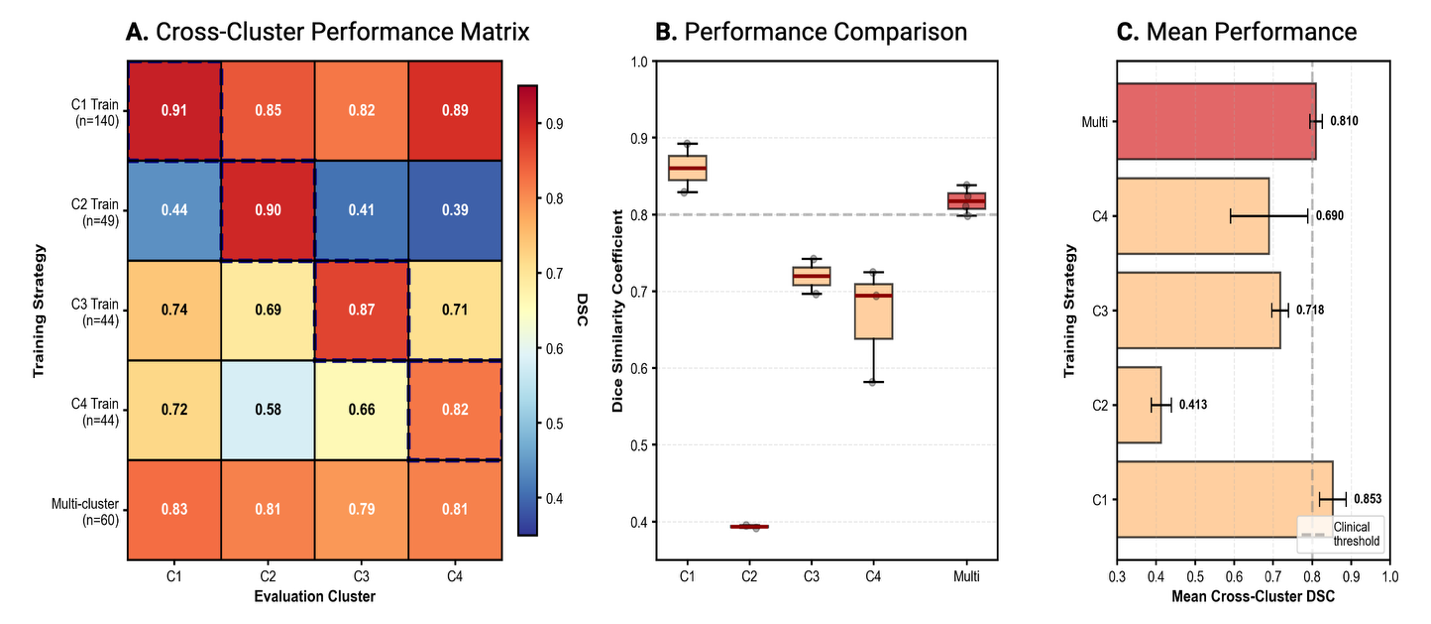

**Figure S2. Comprehensive cross-cluster generalization analysis.** **(A)** Performance matrix (heatmap) showing DSC values for models trained on individual clusters (rows 1-4) or multi-cluster data (row 5) evaluated across all four technical clusters (columns). Color scale: blue (DSC<0.50), yellow (0.50-0.75), red (DSC>0.85). Dashed boxes indicate within-cluster performance. **(B)** Box plots comparing cross-cluster performance distributions for single-cluster (orange) versus multi-cluster (red) training strategies. Gray dashed line: clinical threshold (DSC=0.80). **(C)** Mean cross-cluster DSC values (±SD) for each training strategy. Multi-cluster training achieves consistent performance (0.810±0.016) above the clinical threshold despite using fewer training cases than most single-cluster approaches, demonstrating that technical diversity matters more than raw sample size for generalization.

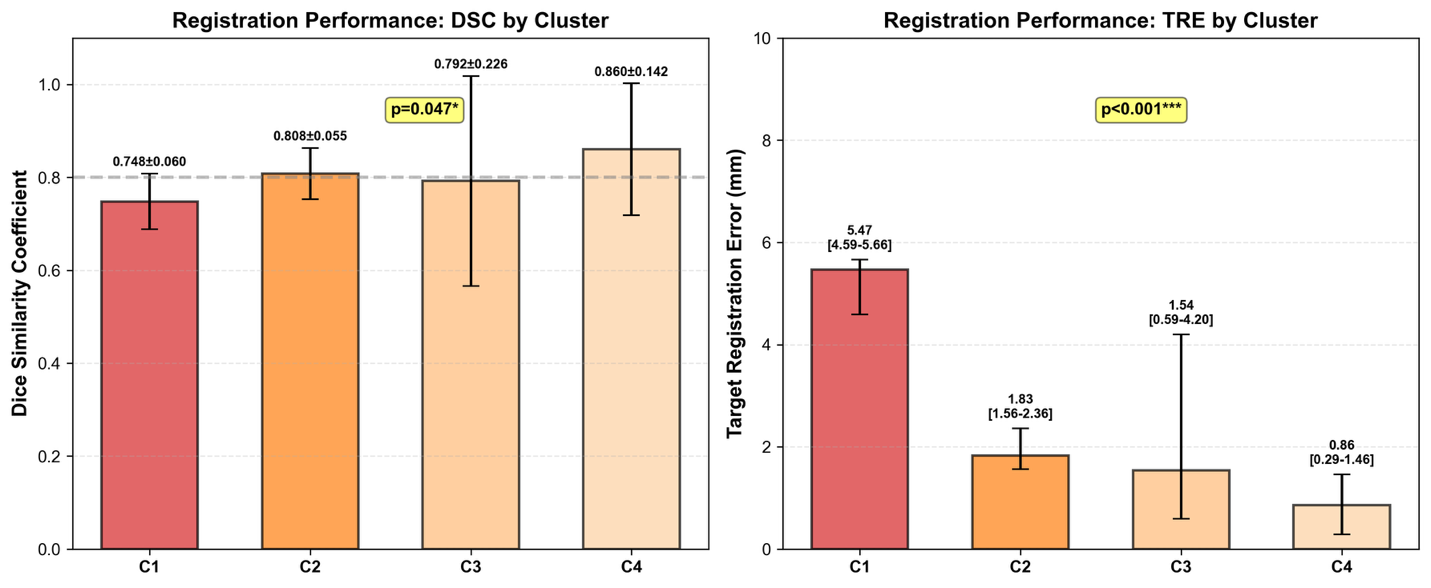

**Figure S3. Registration performance varies significantly across MRQy-defined image quality clusters.** Dice Similarity Coefficient (DSC) across the four MRQy-derived image quality clusters. Bars represent the **mean ± standard deviation**. Target Registration Error (TRE, in millimeters) across the same clusters. Bars represent the **median**, with **interquartile range (IQR)** shown as error bars. Significant differences between clusters are annotated with the corresponding p-values.

**Experiment 2.** Prostate Segmentation Model (M₁): Architecture and Preprocessing

To evaluate the impact of batch effects on prostate gland segmentation, we trained and tested a V-Net deep learning model (M₁) on T2w MRI scans using the MRQy-defined clusters from Experiment 1. Ground truth prostate gland segmentations were manually annotated by a board-certified radiologist (S.A., 9 years of subspecialty training in genitourinary imaging, annual volume >1,500 prostate MRI examinations, fellowship-trained in abdominal and pelvic imaging) using 3D Slicer software.

Two complementary experiments were conducted:

• **Single-cluster training:** The model was trained on individual clusters and evaluated on the remaining clusters to quantify the generalization gap introduced by training on site-specific data.

• **Balanced multi-cluster training:** Fifteen cases were randomly selected from each cluster to create a balanced dataset (n=60 total). The model trained on this subset was then evaluated on the remaining, non-overlapping data from all clusters.

**Preprocessing:** Input images were resampled to a standardized voxel spacing of 0.5×0.5×3.0 mm and resized to 96×96×32 voxels using trilinear interpolation. Images were normalized to the [0,1] range using min-max scaling. Segmentation masks were binarized (threshold=0.5), and data augmentation was applied during training including random flipping along all three axes with 50% probability.

**Architecture and Training:** The V-Net architecture followed a hierarchical encoder-decoder design with five encoding stages, a bottleneck, and five decoding stages. The encoder employed strided convolutions for downsampling with filter numbers progressively increasing (16, 32, 64, 128, 256), and the decoder used transposed convolutions for upsampling with symmetric filter reduction. To improve performance and address potential undertraining concerns, we implemented several architectural enhancements: (1) residual connections at each encoder and decoder stage to facilitate gradient flow, (2) batch normalization layers after each convolution to stabilize training, (3) attention gates in skip connections to focus on relevant features, (4) PReLU activation functions instead of standard ReLU, and (5) spatial dropout (rate=0.3) for regularization. Training used Dice loss to address class imbalance, optimized with Adam (initial learning rate 1e-4). A ReduceLROnPlateau scheduler decreased the learning rate by 0.2 after five epochs without improvement. Models were trained for up to 100 epochs, with early stopping based on validation Dice score convergence. All training was implemented in TensorFlow on NVIDIA RTX A5500 GPUs (24 GB).

**Evaluation:** Performance was assessed using Dice similarity coefficient (DSC), precision, recall, Intersection over Union (IoU), Hausdorff Distance 95th percentile (HD95), and volume ratio. The volume ratio, defined as the predicted-to-ground-truth volume ratio, quantified systematic over- or under-segmentation. Statistical comparisons across clusters were performed using the Kruskal-Wallis test, and p<0.05 was accepted as statistically significant.

**Experiment 3.** Deformable image registration of ADC Maps onto T2w Images: Architecture and Preprocessing

To evaluate the impact of batch effects on deformable image registration, we trained and tested a VoxelMorph-based model (M_2_) using MRQy-defined clusters from Experiment 1. In each case, the T2w image served as the fixed reference due to its superior spatial resolution, while the ADC map was treated as the moving image.

**Registration pipeline:** A two-stage process was employed. First, rigid registration was performed using mutual information with random sampling of 32 voxels to achieve efficient global alignment. This was followed by affine registration, also using mutual information, to account for translation, rotation, scaling, and shearing. This hierarchical approach corrected both global and local misalignments while maintaining computational efficiency. Quality assurance was performed visually, and in nine cases with gross misalignment, the original T2w and ADC pairs were preserved for downstream use to avoid reducing the dataset size.

**Model implementation:** The deformable registration network was implemented in PyTorch/MONAI and trained on NVIDIA RTX A5500 GPUs (24 GB memory).

**Preprocessing:** Images were center-cropped to fractional dimensions (4/8, 4/8, 8/8) along x, y, and z, resized to 64×64×32 voxels using bilinear interpolation, and intensity-normalized by clipping between the 1st and 99th percentiles and scaling to [0,1].

**Augmentation:** To improve generalizability, probabilistic 3D augmentations were applied during training, including random rotations (±10°), translations (±5 voxels), zooms (0.9–1.1×), and flips, each with 50% probability. Identical random seeds ensured paired inputs were transformed consistently.

**Training:** A five-fold cross-validation strategy was used on 277 image pairs. Each fold was trained for up to 100 epochs with early stopping (patience = 15). The AdamW optimizer (lr = 1e-4, weight decay = 0.01) was used, and a ReduceLROnPlateau scheduler halved the learning rate after five stagnant epochs. After cross-validation, the best configuration was retrained on the entire dataset for 100 epochs to maximize data use. A composite loss function guided the training process.

**Loss function:**

$$Total Loss=MSE+\beta(L1+nCC)+\alpha(Gradient Loss)$$

Mean squared error (MSE) served as the primary similarity term to align image structures. To account for intensity variation across modalities, both L1 loss and normalized cross-correlation (nCC) were included and jointly weighted by β = 0.25. A gradient loss term was added to enforce smooth and anatomically plausible deformation fields, weighted by α = 0.05.

**Evaluation:** Model performance was assessed using DSC for volumetric overlap and Target Registration Error (TRE) for spatial accuracy. DSC measured gland overlap between fixed and registered images, while TRE quantified center alignment.
